# Modelling the decadal expansion of West Nile virus in Italy: the role of climatic, anthropogenic, and macroecological drivers

**DOI:** 10.64898/2026.06.17.26355538

**Authors:** Lara Marcolin, Antonino Bella, Martina Del Manso, Ilaria Dorigatti, Patrizio Pezzotti, Piero Poletti, Flavia Riccardo, Moreno Di Marco

## Abstract

**BACKGROUND:** West Nile virus (WNV) is a growing health burden in Italy. Anticipating human infection risk is hampered by the pathogen’s complex ecology, highlighting the need for comprehensive early-warning tools.

**AIM:** We aimed to model municipal-level WNV risk in Italy and characterize its decadal expansion in Italy, providing a comprehensive ecological understanding of viral emergence.

**METHODS:** We applied a machine learning framework to annual human WNV case data from 2014 to 2024. The model integrated a suite of environmental, socio-economic, and macroecological predictors to generate risk projections. We evaluated the model’s performance through multiple validation settings. We also performed an anticipation test for the 2025 epidemic season, using 2024 environmental data to assess the model’s predictive accuracy against observed 2025 human cases.

**RESULTS:** Our model achieved robust performance (True Skill Statistic > 0.4) and captured WNV progressive expansion from 184 predicted positive municipalities in 2014 to 2,012 in 2024 (an 11-fold increase in 11 years). Seasonal minimum temperature was the primary risk driver, followed by monitoring year and population density, indicating active spatial spread. Environmental suitability consistently preceded clinical detection. Municipalities with cases in 2023–2024 exhibited significantly higher predicted suitability during 2018–2022 than those without cases (average risk 0.58 *vs* 0.20). Our model successfully identified emerging risk hotspots along the Adriatic coast and southern Italy before the official human spillover of 2025.

**CONCLUSION:** Embedding macroecological drivers into WNV risk modelling provides an improved understanding of drivers of rapid WNV expansion. Our model enables proactive risk mapping, surveillance efforts, and targeted public health measures.

## 1 Introduction

The expansion of West Nile virus (WNV) poses a major public health threat in Europe, with rapid increase in outbreak frequency and intensity over the last decade [1]. Italy has emerged as a critical hotspot of WNV transmission, consistently reporting the highest number of human neuroinvasive disease cases in the continent [2]. Despite its public health relevance, predicting WNV spillover remains difficult due to its multi-vector, multi-host, transmission system which involves several mosquito vectors and several avian hosts [3]. To monitor this complex zoonosis, Italy has implemented the 2020–2025 National Arbovirus Plan which employs a One Health approach integrating seasonal human, entomological, and avian surveillance to prompt timely preventive and mitigation measures [4]. While this system is essentially designed to track viral circulation, it remains fundamentally reactive, triggering interventions only after human or animal infections have occurred [5]. This reactive paradigm is perpetuated by a critical disconnect between field surveillance and spatial modelling. While empirical monitoring and statistical analyses have consistently identified the role of ecological factors in modulating WNV risk, current models of human cases often fail to incorporate such ecological insights into risk prediction in Italy [6,7].

In fact, these models often operate at coarse spatial resolutions, limiting their utility for local interventions, and lack robust spatio-temporal validation for real-world forecasting [8]. They also rely primarily on climatic and land-use predictors while overlooking key processes underlying WNV outbreaks, including biotic interactions and macroecological drivers regulating viral circulation [9,10]. For instance, specific levels of ecological integrity and avian biodiversity can directly influence the abundance of competent host species, thereby regulating WNV circulation between mosquitoes and wild birds [11,12], yet these factors remain absent from current operational maps.

Here we develop a municipal-level WNV risk model for Italy, which integrates human, climate and macroecological drivers [9]. By combining high-resolution maps of avian biodiversity and ecosystem integrity with climatic and anthropogenic data, we provide a granular characterization of the drivers of WNV across Italian municipalities through time. Our goal is to offer an ecologically grounded basis for targeted surveillance and early-warning interventions by public health and veterinary services.

## 2 Methods

We applied a machine learning framework to predict the presence of WNV cases across Italian municipalities and to characterize the spatio-temporal dynamics of WNV expansion. Our model integrated annual records of human incidence data with climatic, environmental, and socio-economic predictors, and was validated using spatial and temporal cross-validation schemes to ensure robust predictive performance in geographically novel areas and for future projections. Furthermore, we assessed the model’s prospective predictive capacity through an anticipation test for the 2025 epidemic season, using 2024 environmental data to forecast observed 2025 human cases.

### 2.1 West Nile virus cases

In Italy, the surveillance of WNV is coordinated by the National Institute of Health (Istituto Superiore di Sanità) and the Ministry of Health under the National Arbovirus Plan [4]. The system adopts a One Health approach, combining human, veterinary, and entomological monitoring to enable early risk detection (Supplementary Text S1).

Based on this framework, we modelled WNV risk across the 7,896 Italian municipalities (i.e., the smallest local administrative units) over an 11-year period (2014–2024). The response variable was derived from municipal-level annual human incidence data provided by the Italian National Institute of Health [4], which aggregates records of neuroinvasive disease, West Nile fever, and positive blood donors. Although ongoing surveillance programs also monitor viral circulation in mosquitoes and horses [6], we deliberately excluded these data to focus our framework on predicting human risk. Human spillover is intrinsically linked to specific drivers of human exposure and socio-economic dynamics, which are distinct from the ecological factors regulating viral transmission in non-human hosts.

Unlike severe neuroinvasive cases, which are consistently detected and subject to mandatory notification, the detection of milder infections heavily depends on active screening. Incidence data were therefore converted into a binary annual presence/absence indicator to minimise heterogeneities in public health surveillance and reporting capacity across different regions, e.g. driven by differences in public investment, surveillance and control efforts. Municipalities were classified as “presences” in a given year (n = 1,645 overall in the period) if at least one human case was recorded. Municipalities with no records for a given year could be either true absences or unobserved data. To address the inherent risk of reporting bias which could lead to false absences (i.e., unobserved data), we defined “pseudo-absences” as those municipalities (n = 59,284 overall in the period) with no recorded cases in a given year but located in provinces (i.e., NUTS-3) reporting at least one confirmed WNV case for the year. This pseudo-absence selection allowed us to minimise the risk of introducing false absences in our analysis, limiting our dataset to areas with known viral circulation and likely higher surveillance effort.

### 2.2 WNV predictors

To capture the multi-dimensional drivers of WNV risk, we integrated a suite of environmental, socio-economic, and macroecological predictors into our model. We derived climatic variables from the MERIDA dataset (∼7 km resolution) [13]. We calculated seasonal mean minimum and maximum temperatures for the May–October period, reflecting conditions relevant for vector activity and viral replication rates [14]. Growing Degree Days (GDD) were computed for March–October using lower and upper thresholds of 11.5°C and 34.7°C to capture cumulative thermal conditions supporting the development of *Culex pipiens*, a key vector of WNV in Italy [15]. Total precipitation (May–October) was included to represent both larval habitat availability and drought-related amplification dynamics [16]. Climatic variables are available up to 2024, restricting the training period of our model (see section 2.3 for the approach used to address data limitations in operational forecasting).

To explicitly account for vector availability, we integrated habitat suitability for *Cx. pipiens* derived from an ensemble species distribution model integrating environmental and anthropogenic predictors [17]. This model integrates anthropogenic (soil imperviousness, population density) and environmental variables to define the species’ thermal and hydrological limits. These data were aggregated at the municipal level using (i) mean suitability and (ii) the proportion of suitable area calculated by binarizing the original suitability layer. Beyond the determination of habitat suitability, land use also plays a critical role in shaping mosquito population size and WNV transmission [16]. Specifically, irrigated croplands and fragmented forests have been positively associated with WNV incidence across Europe. These landscapes create stable larval habitats and ecotones that facilitate the proximity between avian reservoirs and mosquito vectors. We accounted for cropland cover using annual layers from WEkEO (www.wekeo.copernicus.eu) at 1 km resolution, from which we derived overall cropland proportion in each municipality per year. As cropland data were only available for the period 2017–2023, we interpolated municipal-level cropland values for the missing years (i.e. 2014-2016 and 2024) through generalized linear models using year as a predictor, with a quasibinomial distribution and logit link.

Macroecological factors focus on the distribution of species, their abundance and their ecological functions at large scales, often acting as key regulators of zoonotic disease risk [9]. Here we included key variables connected to the dilution effect hypothesis, where higher avian diversity may reduce infection rates by dispersing blood meals across less competent reservoirs [11]. We accounted for bird species richness derived from high-resolution (1 km) species distribution models [18], averaged at the municipal level. We also accounted for environmental variables representing overall forest cover and ecosystem integrity, which are known to influence zoonotic viral circulation [8,10]. We included the annual percentage of forest cover per municipality from the Global Forest Change dataset [19] with an original resolution of 30 m. To characterize ecological integrity, we included the Biodiversity Intactness Index at 1 km resolution [20], which quantifies the average abundance of native species relative to an undisturbed state. High values suggest a more pristine community assembly, which may be more resilient to the dominance of highly competent synanthropic bird species, such as certain passerines [21], that typically thrive in degraded or simplified environments and drive viral amplification [22].

We included year as a variable in our model, reflecting both the progressive expansion of WNV in Italy and the likely increase in diagnostic sensitivity and surveillance effort over time. Municipality area and log-transformed human population density were also utilized to represent drivers of human exposure to mosquito bites. Population data were obtained from the LandScan Global dataset at a 1 km resolution [23], while municipality area from the national administrative boundaries dataset were obtained by the Italian National Institute of Statistics [24].

All predictors were spatially aggregated to the municipal level using area-weighted mean value of all grid cells falling within each municipality’s administrative boundaries. Before model training, we assessed multicollinearity using pairwise Spearman’s correlation and excluded variables with |r| > 0.7 (Figure S1).

### 2.3 Predictive modelling framework

The model was trained on the complete dataset, comprising 60,929 unique spatio-temporal presence/pseudo-absence observations recorded between 2014 and 2024. We employed a Balanced Random Forest (BRF) algorithm, implemented via the *randomForestSRC* package [25], to address the severe class imbalance in our dataset (i.e. 40 times more pseudo-absences than presences). BRF utilizes a down-sampling strategy [26] where each individual tree is trained on a balanced bootstrap sample created by combining all presence observations with an equal number of randomly selected pseudo-absences. The forest was grown using 1,000 trees, the number of candidate variables randomly selected at each split (mtry) was set to 2, and the minimum terminal node size (nodesize) was set to 5, as determined through hyperparameter tuning. Based on continuous model predictions for each municipality (i.e. probabilities of having WNV cases), we defined a binary classification (i.e. positive *vs* negative municipalities) applying a standard classification threshold of 0.5 to predicted probabilities [27]. We preferred not to derive an optimal threshold (e.g. based on validation performance, see below) to maintain consistency across the different cross-validation schemes and ensure that the performance results from the spatial and temporal tests remained directly comparable.

We evaluated the model’s performance using two validation approaches [28]. First, we performed a spatial block cross-validation using a leave-one-region-out scheme, where all municipalities within a given region were excluded from training and used for independent testing. Only regions with more than 10 municipalities with presences were retained as validation units. This procedure was repeated iteratively across regions to assess model transferability to geographically novel areas. Second, we conducted a temporal out-of-time validation to simulate real-world operational forecasting. We trained the model on a reduced dataset covering the period 2014–2023, and tested its predictive power against 2024 data. This step was necessary to verify that the model remains effective despite year-to-year fluctuations in climatic and ecological drivers. The training dataset consisted of 1,396 presence observations and 54,021 pseudo-absences (a ratio of approximately 1:40), while the 2024 test set included 276 presence observations and 5,263 pseudo-absences. To evaluate the model’s capacity to forecast risk in real-world operational scenarios—where current-year climatic data might not be immediately available—we also conducted an anticipation test for the 2025 epidemic season (Supplementary Text S2). Specifically, we predicted the risk associated with 2025 human cases by employing environmental and climatic covariates from the previous year (2024), while only updating the temporal variable in the model (i.e., setting “year” to “2025”).

Model performance was evaluated using sensitivity (i.e., recall rate), specificity, true skill statistic (TSS, combining sensitivity and specificity) [29], precision, and the F1 score (i.e., combining recall and precision) [30]. To ensure the reliability of these projections, we also performed a Multivariate Environmental Similarity Surfaces analysis [31] using the *dismo* package [32] to identify areas where predictors fall outside the training range, thus highlighting regions where predictions represent an ecological extrapolation and should be interpreted with caution.

Finally, we used the trained model to generate annual risk maps (2014–2024) at the municipal scale based on year-specific predictor values. To visualize the spatio-temporal expansion of WNV, model predictions were compared with the annual distribution of provinces reporting human cases, enabling direct comparison between the model’s predicted WNV risk and the observed spatio-temporal expansion of WNV occurrence in humans across Italy through time.

Statistical inference was performed using the R package *randomForestSRC* [25]. Variable importance was quantified as the mean decrease in model performance following covariate permutation. Partial dependence plots were used to represent the marginal effects of predictors on WNV risk.

## 3 Results

### 3.1 Spatio-temporal trends of WNV human cases

Between 2014 and 2025, Italy experienced marked inter-annual fluctuations in West Nile virus transmission, characterized by years of low-level endemic circulation punctuated by major epidemic outbreaks (Figure 1; Figure S2). During these severe seasons, the spatial extent of viral transmission expanded significantly. As shown in Figure 1, the years 2018, 2022, 2024, and 2025 emerged as the most severe, with the number of affected municipalities peaking at over 400 in 2022 and 2025 (Figure S2a). This geographic expansion directly correlated with sharp peaks in the total number of reported human WNV cases, which exceeded 600 in 2018 and nearly reached 800 in 2025 (Figure S2b). Spatially, while the endemic core of the virus remained concentrated in the Po Valley in northern Italy, the annual occurrence maps clearly illustrate a progressive southward expansion over the study period, with an increasing density of cases affecting central regions, the Adriatic coast, and the major islands.

**Figure 1.**
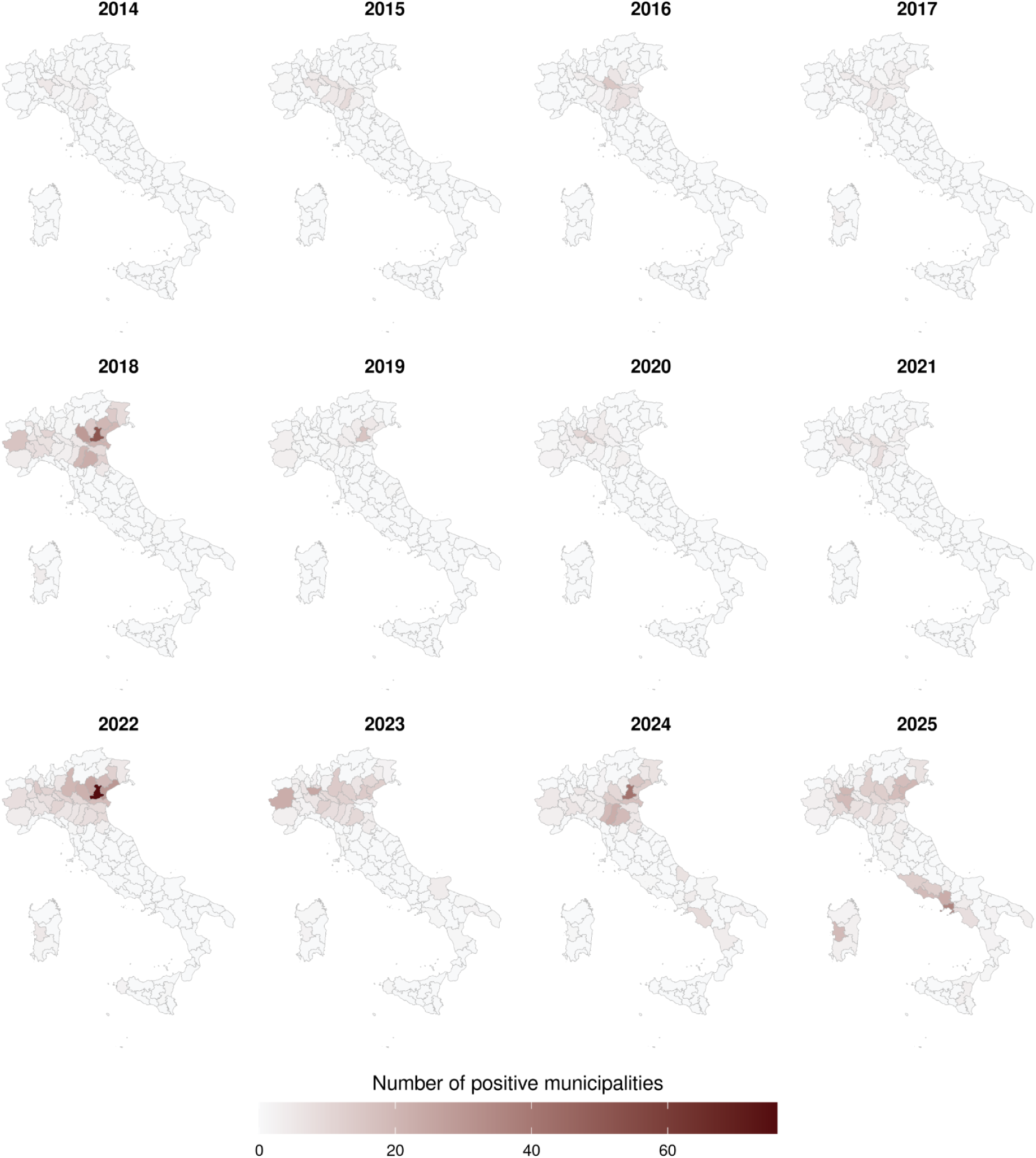
Spatio-temporal distribution of human WNV cases across Italian municipalities (2014–2025). The panel of maps tracks the annual geographical spread and intensity of reported WNV infections. The color gradient indicates the number of positive municipalities at the province level (NUTS 3).

### 3.2 Model performance and spatio-temporal extrapolation

Our modelling framework showed high predictive accuracy and robustness in discriminating the occurrence of human WNV cases across Italian municipalities, being able to effectively address the extreme class imbalance between presences *vs* pseudo-absences (Table 1). The spatial block-validation showed good discriminative ability (TSS>0.4), although regional performance exhibited significant variability (Table S1). Predictive accuracy was higher in northern endemic regions, such as Emilia-Romagna (TSS = 0.54), Veneto (TSS = 0.49), and Friuli-Venezia Giulia (TSS = 0.66), while lower values were observed in regions like Piedmont (0.30) and Sardinia (0.21), likely reflecting local environmental conditions and context-specific transmission dynamics not fully captured by municipality-scale predictors. The temporal validation yielded higher predictive performance, with an average TSS of 0.61 (Table 1) and a high sensitivity (>0.9). This is particularly relevant for public health application, as it enhances the ability to detect the emerging municipalities at risk, while minimising false negatives, a critical requirement for early-warning systems.

**Table 1.**
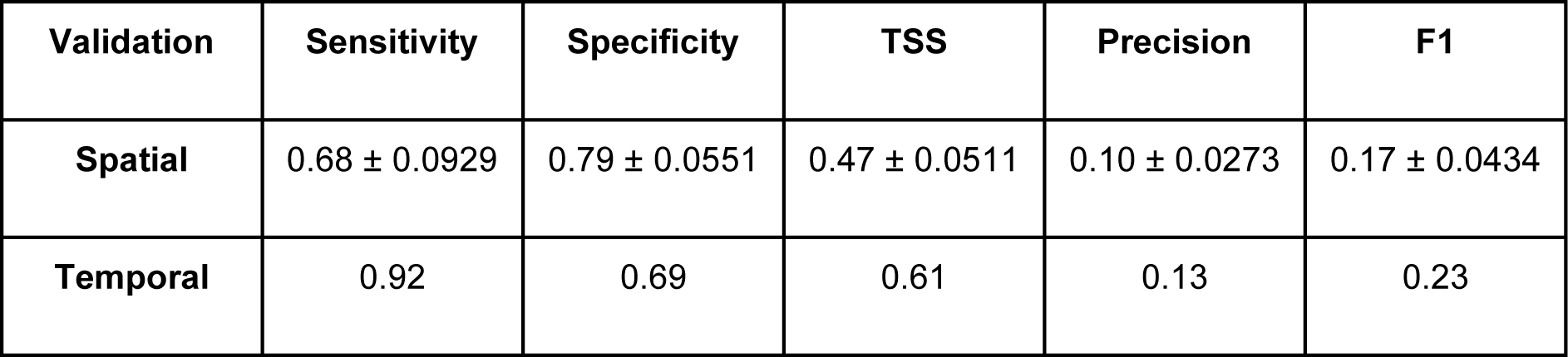
Predictive performance metrics. Summary of the model’s performance using two distinct approaches: the spatial block cross-validation (leave-one-region-out) and the temporal out-of-sample validation (tested on 2024 data). Results for the spatial validation represent the mean values across all validated regions, accompanied by the standard error of the mean.

| Validation | Sensitivity | Specificity | TSS | Precision | F1 |
| --- | --- | --- | --- | --- | --- |
| **Spatial** | 0.68 ± 0.0929 | 0.79 ± 0.0551 | 0.47 ± 0.0511 | 0.10 ± 0.0273 | 0.17 ± 0.0434 |
| **Temporal** | 0.92 | 0.69 | 0.61 | 0.13 | 0.23 |

Values for precision (0.10–0.13) and F1-score (0.17–0.23) were lower across both validation schemes, reflecting the intrinsic difficulty of predicting a rare event in a highly imbalanced dataset, where WNV diagnoses constitute only a small proportion of the total infections. Despite these constraints, performance metrics improved substantially in endemic areas like Emilia Romagna (F1=0.35) and Veneto (F1=0.31), indicating high reliability in areas with consolidated viral circulation.

The sensitivity analysis and anticipation test confirmed that the model maintains high predictive robustness even when relying on climatic data from the previous year (Table S2), further supporting its utility as an operational early-warning tool. In the sensitivity analysis for 2024, using 2023 covariates yielded results nearly identical to those of the standard model employing synchronous variables. Across the national territory, sensitivity remained exceptionally high in both cases (0.92), with matching TSS values (0.66). Within the subset of 65 historically affected provinces, performance was similarly stable (TSS∼0.6).

When applying this anticipatory approach to predict 2025 WNV presence, using covariates from 2024, the model demonstrated robust discriminative ability. At the national scale, the model achieved a sensitivity of 0.71, a specificity of 0.77, and a TSS of 0.48 (F1-score=0.24). Restricting the evaluation to the historical training area increased sensitivity to 0.74 and yielded a slightly improved F1-score (0.25), while maintaining a comparable TSS (0.46).

### 3.3 Key drivers of WNV risk

Analysis of variable importance through permutation identified minimum temperature (May–October) as the most influential predictor of WNV presence (Figure 2). This was followed by human population density and the calendar year. Among the macroecological factors, bird species richness provided a substantial contribution to the model’s performance, confirming that the structure of the avian reservoir community is a fundamental component of the WNV transmission cycle in Italy. Partial dependence plots revealed distinct non-linear relationships and ecological thresholds for the primary risk drivers (Figure 3). The probability of WNV presence showed a sharp, non-linear increase when minimum temperatures reached approximately 15°C, peaking around 20°C. In contrast, cumulative precipitation (May-Oct) shows a relatively flat profile compared to other variables, suggesting that at the municipality scale, total seasonal rainfall has a more marginal effect on human risk than thermal and ecological drivers.

**Figure 2.**
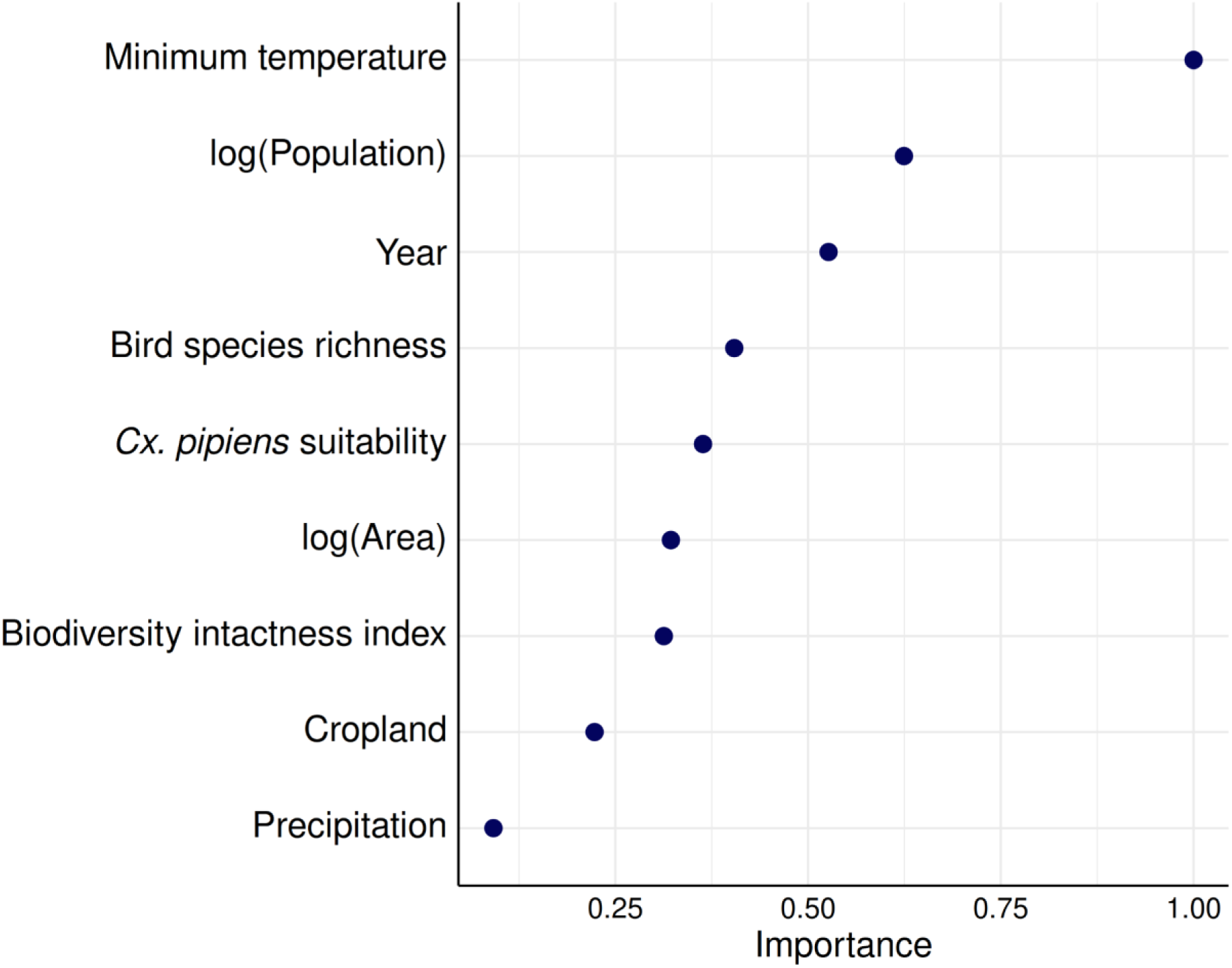
Relative importance of predictors for West Nile virus risk. The plot illustrates the contribution of environmental, socio-economic, and macroecological variables to the BRF model, measured via mean performance reduction following covariate permutation. The variable “*Cx. pipiens* suitability” represents the proportion of suitable habitat for the primary vector within each municipality.

**Figure 3.**
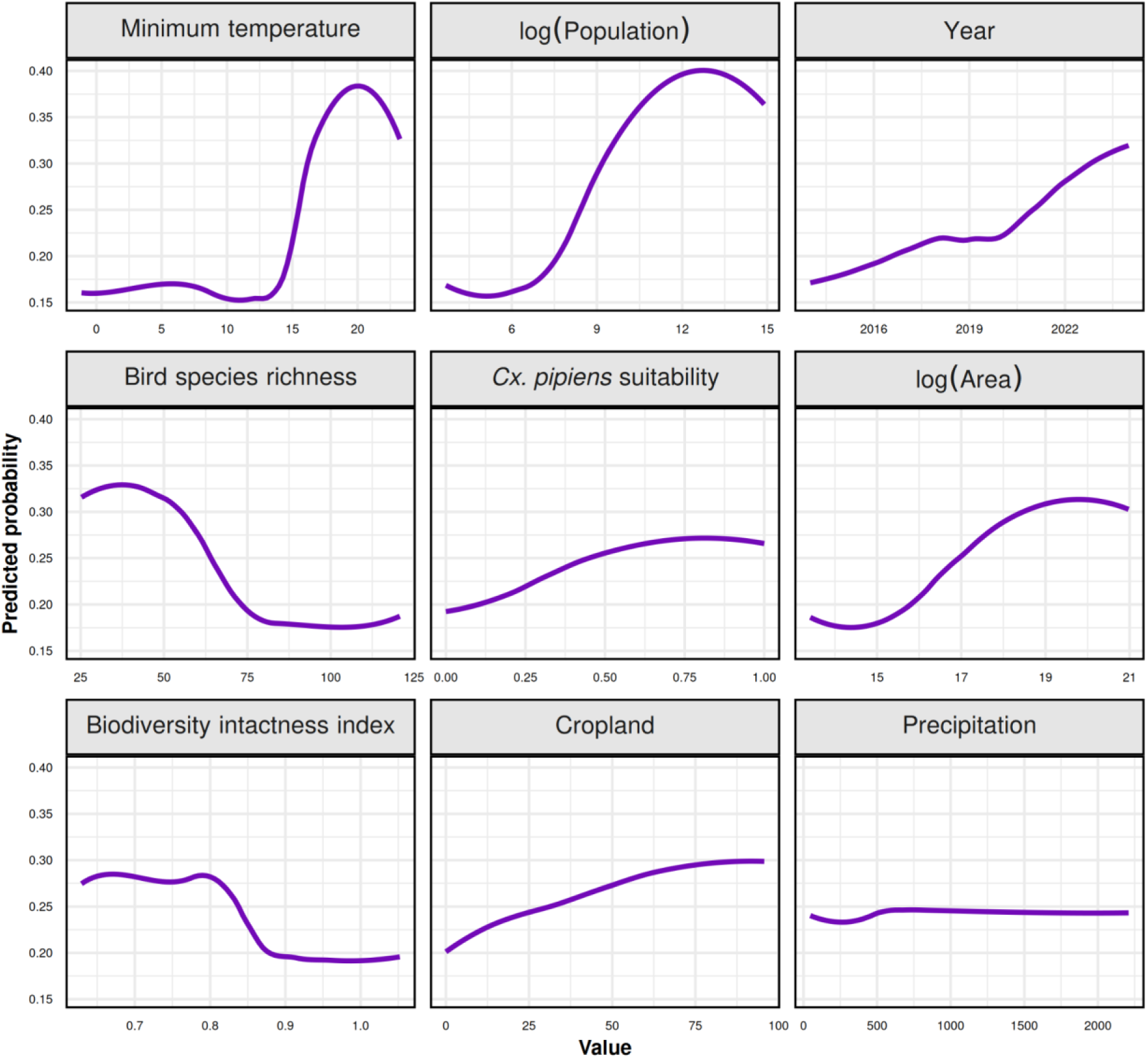
Partial dependence plots. The plots illustrate the marginal effects of individual environmental, socio-economic, and macroecological predictors on the predicted probability of WNV presence at the municipal level. The y-axis represents the predicted probability of risk, while the x-axis shows the range of values for each predictor. The variable “*Cx. pipiens* suitability” represents the proportion of suitable habitat for the primary vector within each municipality.

Bird species richness shows that the risk is highest at low levels of diversity and drops significantly as the number of species increases. Similarly, the Biodiversity Intactness Index plot shows that WNV risk is higher in degraded ecosystems (lower values) and decreases as ecosystem integrity improves, with a marked drop after the 0.85 threshold. Taken together these two patterns confirm a dilution effect for WNV at national scale. Finally, the calendar year shows a steady, almost linear upward trend, reflecting the progressive temporal expansion of WNV in Italy. Instead, human population density shows that risk sharply increases with density before reaching a plateau.

### 3.4 Risk mapping through space and time

We projected WNV risk for the year 2024 at 1 km spatial resolution (Figure 4), and identified high-risk areas primarily in the Po Valley, as well as emerging hotspots along the Adriatic coast and in southern regions that have historically reported fewer cases.

**Figure 4.**
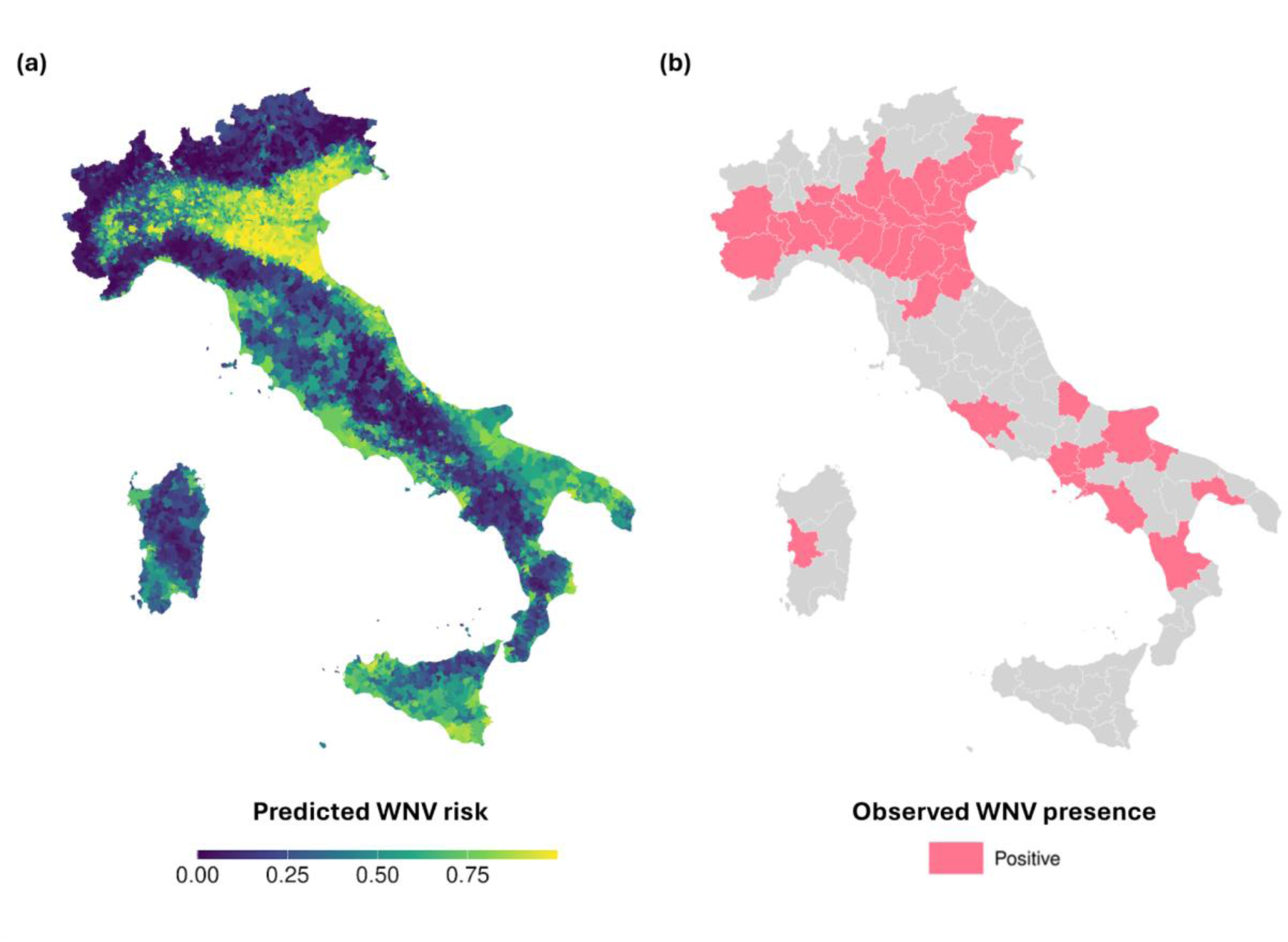
Predicted and observed West Nile virus circulation in Italy for the 2024 epidemic season. (a) Spatial distribution of human transmission risk at the municipal scale, based on the 2024 environmental and climatic conditions. The color gradient represents the predicted probability of WNV risk, with yellow indicating high-risk zones and dark purple representing low-risk areas. (b) Observed WNV presence at the provincial level during 2024; provinces are classified as positive (pink) if at least one human case was detected.

The multi-annual risk projections revealed a clear expansion of high-risk areas in Italy, moving from the historically endemic areas of the Po Valley towards the Adriatic coast and central-southern Italy (Figure 5). The chronological overlay of provinces with recorded human cases showed a strong correspondence between expansion of suitable conditions for WNV occurrence and expansion of observed cases.

**Figure 5.**
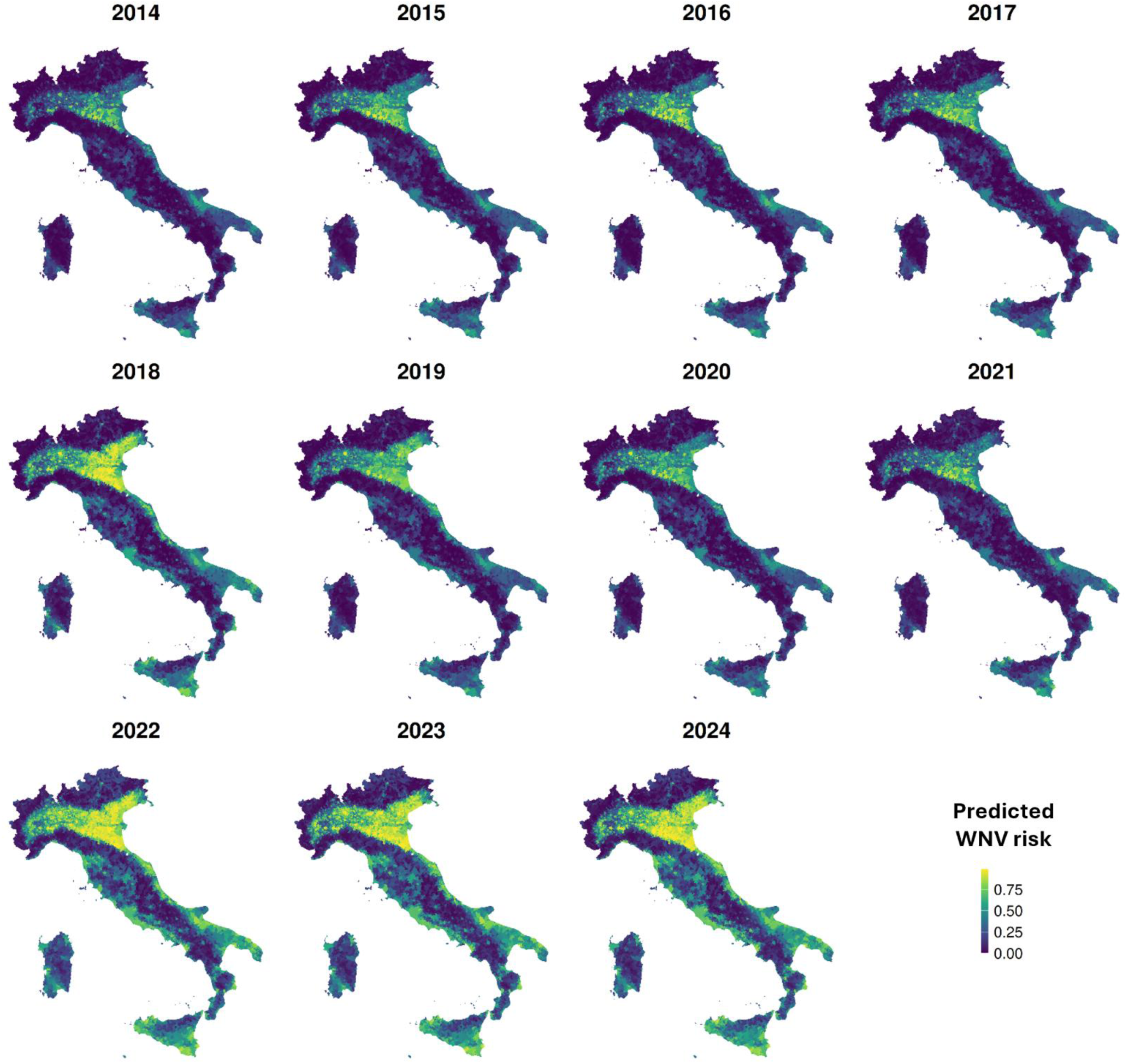
Spatio-temporal dynamics of predicted WNV risk and historical expansion. The panel illustrates the annual progression of predicted risk maps (2014–2024) to represent the decadal trend. The color scale indicates the predicted probability of WNV risk at the municipal scale.

Notably, the model identified areas of high environmental suitability before the first human cases were detected in certain provinces, reflecting the virus’s progressive spread and the utility of our modelling tool. For example, the model identified a marked increase in WNV risk during 2018 and 2022, effectively anticipating the spatial spread in regions that recorded their first human cases in subsequent years [33]. Indeed, the average predicted risk for 2018-2022 was three times higher in municipalities that became positive in 2023-2024 than in those that did not (0.58 *vs* 0.20).

## 4 Discussion

We developed a municipal-level macroecological risk model that effectively captured the spatio-temporal dynamics of WNV circulation in Italy between 2014 and 2024. Our results showed a clear decadal expansion of WNV suitability from the endemic core of the Po Valley toward the Adriatic coast and southern regions, which was followed by a congruent expansion in human cases. The temporal analysis highlighted a “facilitation effect”, wherein environmental suitability systematically precedes pathogen geographical expansion. The strong temporal validation results (TSS: 0.61; Sensitivity: 0.92) indicates that WNV spread follows predictable environmental and climatic patterns over time. Our spatial validation provides further insight into this expansion by revealing a clear biogeographical gradient in the model’s predictive behaviour. In the endemic Po Valley, where viral circulation is fully established [2], environmental suitability consistently translated into clinical cases. This is reflected in high F1-scores in regions like Emilia-Romagna and Veneto, indicating strong model reliability. Conversely, in expansion areas such as Apulia and Campania, we observed high sensitivity but low precision, limiting F1-scores. This reflects low pathogen prevalence and local heterogeneity or stochastic introduction dynamics that operate below the municipal resolution. Furthermore, the low precision observed in expansion areas aligns with recent evidence indicating that the incidence of reported clinical cases is heavily influenced by heterogeneous surveillance sensitivities and does not necessarily reflect the true underlying risk of WNV infection [34]. However, from a public health perspective, this pattern exemplifies our model’s early-warning capacity. When combined with the temporal facilitation effect, high sensitivity in southern regions suggests that identified expansion frontiers represent concrete risks. By highlighting territories that are biologically primed for spillover, before sustained incidence occurs, our framework provides actionable lead time which might anticipate the presence of human cases by one year [1,33]. This is well-illustrated by the 2025 WNV outbreak in Lazio, where our model’s high suitability scores successfully highlighted the emergence of human spillover in this previously under-reported area well ahead of the clinical detection of cases [35].

A key advancement of our model is the integration of macroecological variables as primary drivers of WNV risk. Avian species richness and the Biodiversity Intactness Index negative correlated with risk, supporting the dilution effect hypothesis already observed for WNV [11,12]. Higher avian richness acts as a buffer against spillover, where a complex bird community includes several poorly competent WNV hosts which reduce virus circulation [36]. Instead, degraded ecosystems tend to support simplified bird communities, typically dominated by competent viral amplifiers [22,34]. In accordance with global patterns [10], we found that high levels of ecological integrity act as a protective barrier against viral circulation. Alongside novel ecological insights, our model confirms the primary role of established climatic and socioeconomic drivers. Minimum temperature emerged as a critical biological predictor, influencing mosquito survival and accelerating viral replication [14]. Calendar year acts as a macro-epidemiological proxy, reflecting both pathogen expansion and increased surveillance sensitivity over time. Human population density also plays a key role, not only by increasing susceptible hosts but by representing human-modified environments preferred by *Cx. pipiens* [17] and synanthropic avian amplifiers [22]. In fact, the suitability for *Cx. pipiens* was positively associated with WNV risk, confirming that suitable conditions for the primary vector are necessary, though not sufficient, for human spillover. Instead, cumulative precipitation had a limited effect at the municipal scale, likely because vector breeding sites depend on highly localized anthropogenic factors—such as drainage systems and water management—not captured by coarse rainfall data [17].

Our anticipation and sensitivity analyses highlight the operational value of our framework for early warning. Predicting 2024 WNV cases using covariates from the preceding year resulted in negligible performance loss compared to using synchronous data. Similarly, our 2025 anticipation test yielded solid predictive accuracy relying entirely on 2024 covariates. This consistency suggests that key drivers of WNV suitability, such as minimum temperature, ecosystem integrity, and avian biodiversity, provide a stable risk baseline that is resilient to inter-annual meteorological fluctuations. Consequently, public health authorities do not need to wait for real-time climatic data to identify emerging hotspots. Proactive interventions, such as vector control and blood-donation screening, can be spatially prioritized well in advance of the epidemic season based on the previous year’s environmental conditions. Our findings are consistent with recent spatio-temporal modelling in Spain, which also successfully utilized one-year lagged predictors to anticipate WNV risk [37]. Our findings open a new operational frontier for WNV control. While focused on Italy, our modelling framework provides a scalable blueprint for European-scale WNV surveillance. Historically, surveillance strategies for WNV in Europe have been predominantly reactive, detecting and containing the virus only after human or animal infections have already occurred. Our findings demonstrated that this paradigm can and should be shifted. By explicitly incorporating macroecological drivers alongside climatic and anthropogenic correlates of WNV, our model contributes to an improved understanding and anticipation capacity of WNV risk. By decoupling risk identification from clinical surveillance intensity, the model enables targeted preventive measures in areas not yet affected but ecologically suitable. This ability to predict expansion frontiers means that the model developed in this study could be utilized as a proactive epidemic tool. We hypothesise that the ecological underpinning behind this tool may be applicable to other vector-borne zoonoses beyond WNV in future work: if emerging pathogens track environmental suitability, macroecological predictions could help identify future hotspots for multiple vector-borne diseases. Integrating such tools into European surveillance systems would enhance preparedness and mitigation capacity. By showing the dilution effect exerted by bird species richness and ecological integrity over WNV risk, our study highlights that biodiversity conservation and ecological monitoring are not merely environmental goals, but foundational strategies to prevent the emergence and spread of zoonotic diseases [38].

## Acknowledgements

This research was supported by EU funding within the NextGeneration EU-MUR PNRR Extended Partnership initiative on emerging infectious diseases (project no. PE00000007, INF-ACT Spoke4) and by the Italian Ministry of University and Research (MUR), under the Fondo Italiano per la Scienza (FIS) - FIS-2023-03158 (CUP: B53C25003010001).

## 5 Data availability

Aggregated data supporting reported results are available upon request to Istituto Superiore di Sanità. Individual-level data are unavailable due to privacy restrictions.

