## Supplementary Material for "Modelling the decadal expansion of West Nile virus in Italy: the role of climatic, anthropogenic, and macroecological drivers"

**Supplementary Text S1: West Nile and Usutu virus surveillance in Italy**

The surveillance of West Nile virus (WNV) and Usutu virus (USUV) in Italy is coordinated by the National Institute of Health (Istituto Superiore di Sanità) in collaboration with the Ministry of Health. It is organized according to the National Arbovirus Plan (2020–2025) [1] and is based on an integrated One Health approach.

The system encompasses human, veterinary, and entomological surveillance. Human surveillance is primarily active during the vector season (May to November) and involves the reporting of suspected or confirmed cases, which are diagnosed through molecular or serological tests.

Animal surveillance plays a critical early-warning role: wild birds (the main reservoirs) and equids are monitored through active and passive systems, allowing for the early detection of viral circulation, often before the appearance of human cases. Concurrently, entomological surveillance relies on the capture and analysis of mosquitoes belonging to the *Culex* genus to detect the presence of the virus in local vectors.

When viral circulation is detected, risk-proportional response measures are implemented. These include the strengthening of local surveillance, targeted vector control interventions, and, critically, safety measures for blood, organ, and tissue donations to prevent transfusion- and transplant-related transmission.

Overall, the system is characterized by the continuous integration of data from humans, animals, and vectors. This framework enables early risk detection and timely responses, with the ultimate objective of preventing severe human cases and limiting the geographical spread of the virus.

**Supplementary Text S2: Sensitivity analysis for the anticipatory approach**

To measure the robustness of the anticipatory approach and quantify any potential loss in predictive performance, we performed a sensitivity analysis applying the same rationale to the 2024 cases (Table S2). For this sensitivity analysis, the model was trained exclusively on historical data spanning from 2014 to 2023. We then validated the 2024 diagnoses using the 2023 covariates and compared these results with those obtained from the standard model trained on the synchronized 2024 covariates. Performance metrics for both analyses were calculated across two different spatial extents: the entire national territory and the specific training area, which comprises the 65 provinces with at least one historically recorded human case

**Table S1. Regional predictive performance from spatial block cross-validation.** The table reports the model's ability to generalize to geographically novel areas using a leave-one-region-out approach. Performance metrics are shown for each administrative region used as a validation unit.

| Region | Presences | Pseudo-absences | Sensitivity | Specificity | TSS | Precision | F1 |
|---|---|---|---|---|---|---|---|
| Piedmont | 180 | 12800 | 0.33 | 0.97 | 0.30 | 0.12 | 0.18 |
| Lombardy | 370 | 15305 | 0.69 | 0.82 | 0.51 | 0.08 | 0.15 |
| Veneto | 580 | 5613 | 0.89 | 0.59 | 0.49 | 0.19 | 0.31 |
| Friuli V.G. | 61 | 2238 | 0.80 | 0.86 | 0.66 | 0.14 | 0.23 |
| Emilia Romagna | 379 | 3251 | 0.91 | 0.63 | 0.54 | 0.22 | 0.35 |
| Campania | 16 | 4681 | 0.63 | 0.88 | 0.51 | 0.02 | 0.04 |
| Apulia | 12 | 2584 | 0.91 | 0.63 | 0.54 | 0.01 | 0.02 |
| Sardinia | 17 | 3118 | 0.24 | 0.97 | 0.21 | 0.05 | 0.07 |

**Table S2. Predictive performance in early-warning scenarios.** The table reports the model's ability to predict human WNV incidence using synchronous climatic and environmental covariates versus covariates of the previous year. Results detail both the sensitivity analysis for the 2024 season—which directly compares synchronous and lagged predictors—and the anticipation test for the 2025 season. Performance metrics are evaluated across two spatial extents: the entire Italian territory ("National") and a focal subset restricted to the 65 provinces with at least one historically recorded human case ("Historically affected provinces").

| Evaluation year | Covariate year | Spatial extent | Sensitivity | Specificity | TSS | Precision | F1 |
|---|---|---|---|---|---|---|---|
| 2025 | 2024 (1-year lag) | Historically affected provinces | 0.74 | 0.72 | 0.46 | 0.15 | 0.25 |
| 2025 | 2024 (1-year lag) | National | 0.71 | 0.77 | 0.48 | 0.14 | 0.24 |
| 2024 | 2024 (Synchronous) | Historically affected provinces | 0.92 | 0.69 | 0.61 | 0.13 | 0.23 |
| 2024 | 2024 (Synchronous) | National | 0.92 | 0.74 | 0.66 | 0.11 | 0.20 |
| 2024 | 2023 (1-year lag) | Historically affected provinces | 0.92 | 0.68 | 0.60 | 0.13 | 0.23 |
| 2024 | 2023 (1-year lag) | National | 0.92 | 0.74 | 0.66 | 0.11 | 0.20 |

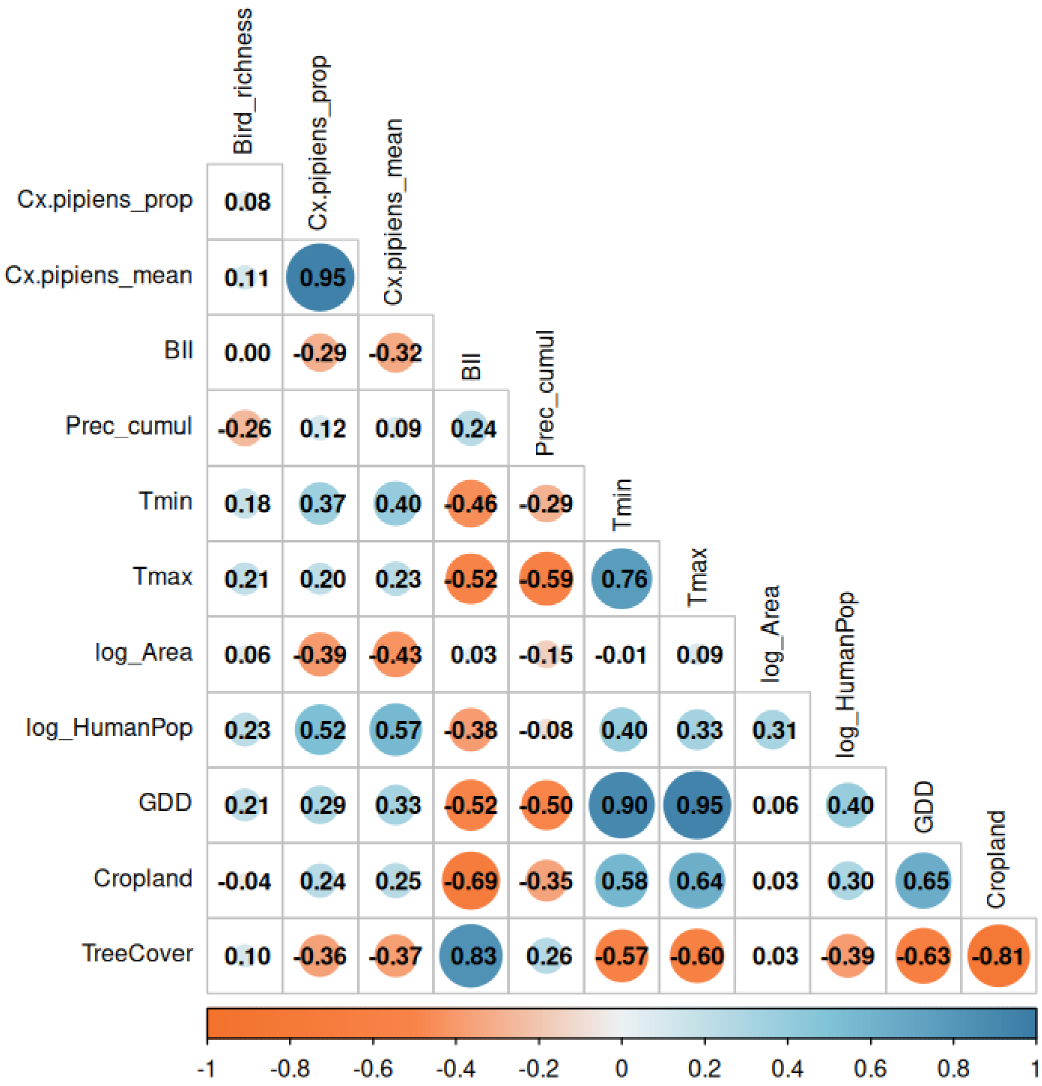


**Figure S1. Spearman's rank correlation matrix of municipal-level predictors.** The correlogram displays the pairwise correlation coefficients among the initial suite of environmental, socio-economic, and macroecological variables.

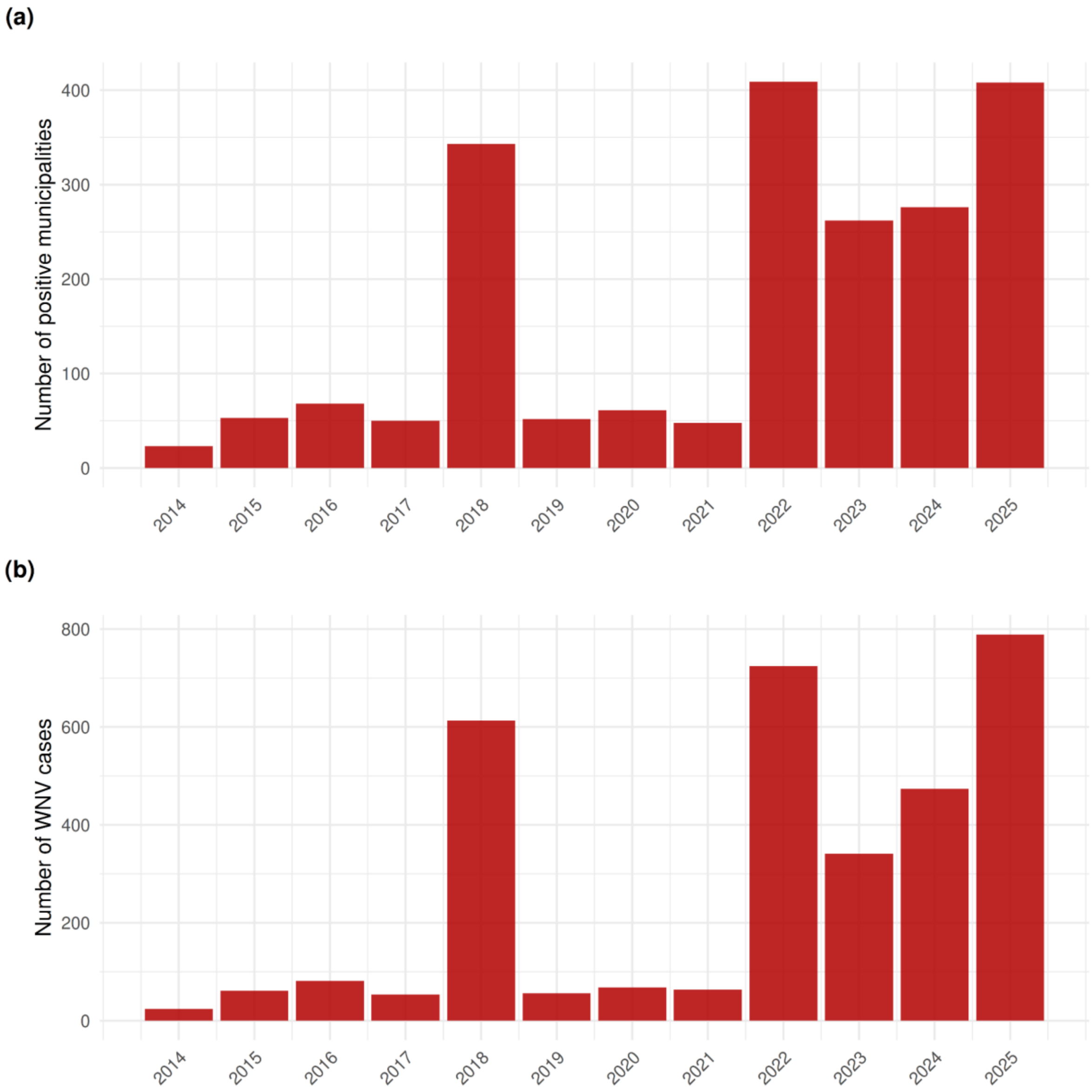


**Figure S2. Inter-annual variability of human WNV transmission in Italy (2014–2025).** The bar charts illustrate the temporal dynamics of the infection over the 12-year study period. Panel (a) shows the total number of Italian municipalities reporting at least one human WNV case per year, reflecting the geographical distribution of the outbreaks. Panel (b) displays the total number of reported human WNV cases per year.

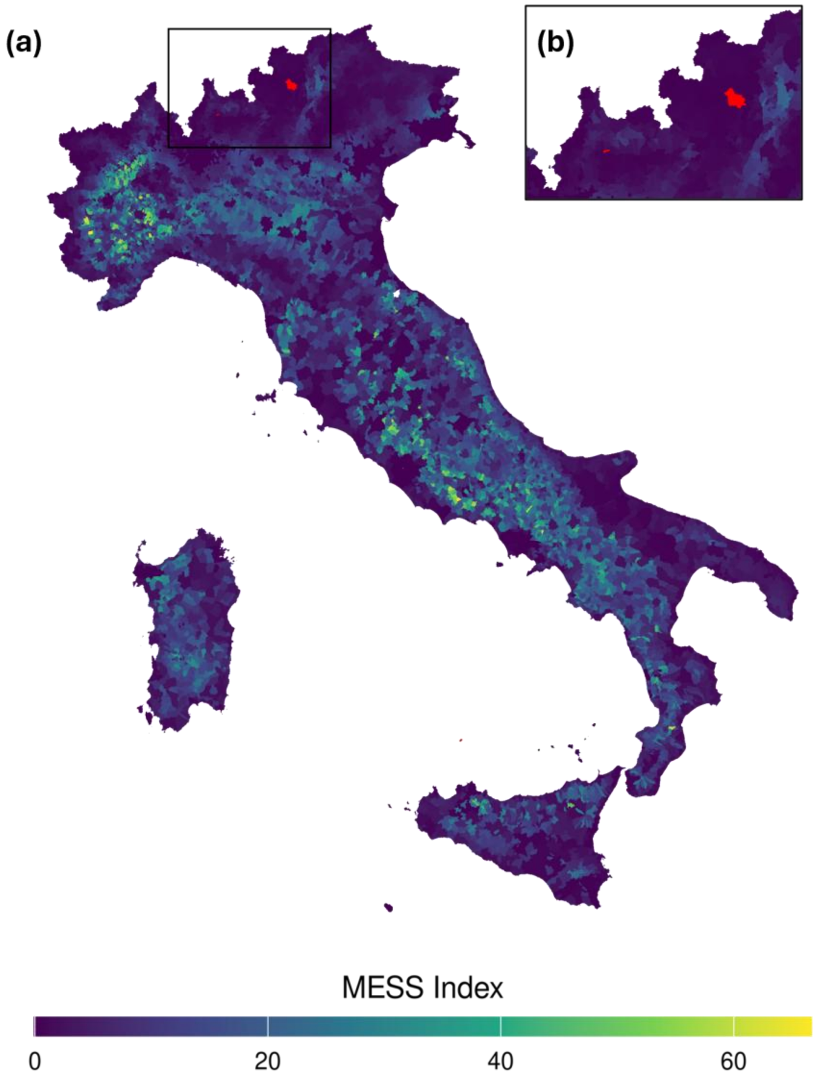


**Figure S3. Multivariate Environmental Similarity Surfaces (MESS) analysis for 2024 risk projections.** The map identifies areas of environmental novelty by comparing the predictor values used for the 2024 projections with the range of values observed during the model training period (2014–2024). Panel (a) shows the MESS index across the entire Italian territory. Positive MESS values (purple to yellow) indicate areas where the environmental conditions for 2024 are within the range of the training data, confirming that risk predictions in these regions are statistically grounded in observed relationships. Conversely, negative MESS values (red) highlight areas of environmental extrapolation, where at least one predictor takes a value outside the training range. Areas with negative values, indicating environmental extrapolation, were primarily confined to high-altitude Alpine regions. Panel (b) provides an inset of the areas in red to better visualize these zones of environmental novelty, where at least one predictor takes a value outside the training range.
